# Association of maternal air pollution exposure and infant lung function is modified by genetic propensity to oxidative stress

**DOI:** 10.1101/2023.10.13.23296994

**Authors:** Dwan Vilcins, Wen Ray Lee, Cindy Pham, Sam Tanner, Luke D. Knibbs, Dave Burgner, Tamara L Blake, Toby Mansell, Anne-Louise Ponsonby, Peter D Sly, Barwon Infant Study Investigator group

**Affiliations:** Child Health Research Centre, The University of Queensland, South Brisbane, QLD 4101; Murdoch Children’s Research Institute, Royal Children’s Hospital, Parkville VIC 3052; Department of Paediatrics, University of Melbourne, Parkville VIC 3052; Florey Institute, The University of Melbourne, Parkville VIC 3052; School of Public Health, The University of Sydney, NSW 2006, Australia; Public Health Research Analytics and Methods for Evidence, Public Health Unit, Sydney Local Health District, Camperdown, NSW 2050, Australia; Melbourne School of Population and Global Health, University of Melbourne, Parkville, VIC 3052, Australia

**Keywords:** Air pollutants, lung function, oxidative stress, environmental health, children’s health, nitrogen dioxide

## Abstract

**Introduction:** The association between air pollution and poor respiratory health outcomes is well established, however less is known about the biological mechanisms, especially in early life. Children are particularly at risk from air pollution, especially during the prenatal period as their organs and systems are still undergoing crucial development. Therefore, our study aims to investigate if maternal exposure to air pollution during pregnancy is associated with oxidative stress (OS) and inflammation in pregnancy or infant lung function at 4 weeks of age, and the extent to which the association is modified by an infant’s genetic risk of OS.

**Methods:** The Barwon Infant Study (BIS) is a longitudinal study of Australian children from the region of Geelong, Victoria. A total of 314 infants had available lung function and maternal OS markers. Exposure to annual air pollutants (NO_2_ and PM_2.5_) were estimated using validated, satellite-based, land-use regression models. Infant lung function was measured by multiple-breath washout, and the ratio of peak tidal expiratory flow over expiratory time was calculated at 4 weeks of age. An inflammation biomarker, glycoprotein acetyls (GlycA), was measured in maternal (36 weeks) and cord blood, and oxidative stress (OS) biomarkers, 8-hydroxyguanine (8-OHGua) and 8-hydroxy-2’-deoxyguanosine (8-OHdG) were measured in maternal urine at 28 weeks. A genetic pathway score for OS (gPFS^ox^) was calculated for each infant participant in the BIS cohort, and high risk defined as score >8. Linear regression was used to explore the association of maternal air pollution exposure with infant lung function, and potential modification by OS genotype was tested through use of interaction terms and other methods.

**Results:** There was no evidence of a relationship between maternal exposure to air pollution and infant lung function in the whole population. We did not find an association between air pollution and GlycA or OS during pregnancy. We found evidence of an association between NO_2_ and lower in functional residual capacity (FRC) for children with a high genetic risk of OS (β=-5.3 mls, 95% CI (−9.3, -1.3), p=0.01). We also found that when NO_2_ was considered in tertiles, the highest tertile of NO_2_ was associated with increase in lung clearance index (LCI) (β=0.46 turnovers, (95% CI 0.10, 0.82), p=0.01) in children with a genetic propensity to OS.

**Conclusion:** Our study found that high prenatal levels of exposure to ambient NO_2_ levels is associated with lower FRC and higher LCI in infants with a genetic propensity to oxidative stress. There was no relationship between maternal exposure to air pollution with maternal and cord blood inflammation or OS biomarkers.

## 1 Introduction

The association of air pollution with respiratory health outcomes is now well established in adult populations, but more work is required to assess risk to children’s health. Air pollution has been associated with increased mortality (1) and increased hospitalisations due to respiratory disease (2) across all stages of life. Children are particularly at risk from air pollution, including during the prenatal period. Maternal exposure to air pollution is associated with adverse child health outcomes, including some adverse respiratory outcomes (3-5), congenital heart defects (6), preterm birth (7) and low birthweight (8). Previous work has shown that maternal exposure to air pollution is associated with infant (4-5 weeks of age) respiratory changes such as poorer lung function (9) lower minute ventilation and higher exhaled nitric oxide (10). Previous evidence consistently demonstrates a strong association between air pollution with poor respiratory health outcomes. However, there are few studies that have investigated the underlying mechanisms of this association, especially in early life (10, 11). Oxidative stress (OS) and inflammation pathways are emerging as key mediators, where exposure to air pollution induces inflammation and/or OS, which then causes damage on a cellular level, leading to poorer lung health (11). The data exploring these processes as mediators between air pollution and children’s health outcomes are scarce. Previous work found higher levels NO_2_ exposure in the prenatal period was associated with an increase in exhaled nitric oxide in infants, which is indicative of airway inflammation (10). A prospective study of asthmatic children in Mexico City found that an OS biomarker, malondialdehyde, in expired breath condensate was associated with exposure to ozone, PM_2.5,_ and presence of heavy vehicles, suggesting that children exposed to high levels of air pollution have higher OS responses (11). In a cross-sectional analysis of Mexican school children, exposure to air pollution increased circulating biomarkers of OS (in serum; conjugated dienes, lipo-hydroperoxides, malondialdehyde, protein carbonylation) in school aged children (7-12 years) (12). A case-control study of Hungarian children revealed an increased risk of infection-induced asthma exacerbation for those children who were more heavily exposed to traffic-related air pollution exposure and had polymorphisms in the NFE2L2 gene, which has a role in regulation of oxidative stress (13).

To the best of our knowledge, only two previous studies have explored the association of maternal air pollution exposure with lung function in infants, and neither have sought to test underlying biological pathways of this association. Further, we could not identify any previous studies that explored maternal exposure to air pollution during pregnancy and OS or inflammation biomarkers. To address this evidence gap, our study aims to investigate if maternal exposure to air pollution during pregnancy is associated with infant lung function at 4 weeks of age and the extent to which this association is modified by a child’s genetic risk of OS. Further, we seek to understand if air pollution exposure is associated with OS biomarkers in mothers, or an inflammation marker in maternal blood during pregnancy or cord blood at birth.

## 2 Methods

### 2.1 Cohort details

The Barwon Infant Study (BIS) is a longitudinal study of Australian children from the region of Geelong, Victoria. The cohort has several major objectives including BIS respiratory, which seeks to investigate factors influencing respiratory health in children. Women were recruited from two major hospitals in the Barwon region and eligible women were invited to participate in the study at a routine antenatal appointment occurring around 15 weeks of gestation. Recruitment occurred between 2010-2013, with ongoing 2-year follow up. A total of 1,064 women who delivered 1,074 infants (10 sets of twins) were enrolled into the study. Participants completed comprehensive questionnaires at 28 weeks antenatal, birth and 1 month of age, which was supplemented with a physical review at each time point. At the 1 month review, participants were invited to undergo multiple breath washout (MBW) testing. The 1 month review was completed by 982 infants and MBW testing was attempted in 570 infants whose parents consented to the test, did not have any respiratory illness, and fell asleep during the test attempt (14). Infants with two or more technically acceptable tests were included in this analysis (*n=*314). The sample size of infants with lung function and maternal OS markers was 257. The study was approved by the Barwon Health Human Research Ethics Committee (HREC 10/24) and written informed consent was obtained from the participating families. Further details on the cohort have been published previously (15).

### 2.2 Air pollutants

We estimated maternal prenatal air pollution exposure using two satellite-based, land-use regression models (Sat-LUR) (16) that we developed and validated and are described in previous studies (17) (18). The Sat-LURs were developed for annual mean concentrations of nitrogen dioxide (NO_2_) and particulate matter ≤ 2.5 μm (PM_2.5_). The indicative spatial resolution of the models is up to 100 m in urban areas and up to 500 m in rural and remote areas (19). NO_2_ is the most spatially heterogeneous component of traffic related air pollution (TRAP) and exhibits strong near-traffic gradients; it is therefore a frequently used proxy for TRAP mixtures. Both Sat-LUR models were developed with satellite, land use and other spatial predictors. The NO_2_ model captured between 66% and 81% of variability in measured annual NO_2_ concentrations, respectively, with RMSE of 1.4 to 2 ppb over the time span of the cohort. The PM_2.5_ model captured between 52% and 63% of variability in PM2.5, with an RMSE of 1 to 1.2 μg/m^3^ (20). Participant’s residential addresses during pregnancy were geocoded to 6 decimal places. These were matched with the air quality data to give estimates of PM_2.5_ and NO_2_ for each participant in the prenatal period, at birth and at 1 month of age. Where participants changed address between follow-ups, their estimated exposure at each property was averaged.

### 2.3 Infant lung function

Infant lung function was measured by multiple-breath washout (MBW) with sulphur hexafluoride (SF_6_) (14) at 4 weeks of age during natural sleep (21). MBW gives indices of lung size (functional residual capacity, FRC) and ventilation distribution (lung clearance index, LCI). LCI provides an early indication of airway obstruction. Tidal breathing was recorded prior to MBW and the ratio of peak tidal expiratory flow over expiratory time (tPTEF/tE) was calculated. A shortening of tPTEF/tE ratio has been used to indicate airway obstruction (22). Two records were excluded from the analysis on the basis of implausible lung function values (FRC >6 L; LCI <1 lung volume turnovers). As previously stated, there was *n*=314 children with data for FRC and LCI, however due to the nature of the test we had a slightly higher sample size for tPTEF/tE (*n*=355).

### 2.4 Inflammation

GlycA is a composite NMR metabolomic marker of glycosylated acute phase proteins (23). Previous work in this cohort have shown that GlycA is a superior measure of cumulative inflammation at a younger age than high-sensitivity C reactive protein (24, 25). GlycA (mmol/L) was measured in maternal (36 weeks) and cord serum (birth) by nuclear magnetic resonance (NMR) (Nightingale Health, Helsinki, Finland). We tested the associations of air pollution with both maternal GlycA during pregnancy and cord blood GlycA in this study.

### 2.5 Oxidative stress biomarkers and gene pathway function score

Maternal urinary samples at 36 weeks of gestation were collected and processed within 24 hours and stored at –80°C until analysis. Urinary OS biomarkers, 8-hydroxyguanine (8-OHGua) and 8-hydroxy-2’-deoxyguanosine (8-OHdG), were quantified using liquid chromatography-mass spectroscopy (LC-MS/MS) performed by the Australian National Phenome Centre (Perth, WA, Australia). Briefly, chromatographic separation was performed using an ExionLC_TM_ system (SCIEX; Framingham, MA, USA), reversed-phase separation used a Kinetex C_8_ 2.6 μm 2.1 × 150 mm column (Phenomenex; Lane Cove West, NSW, Australia) at 40°C, and mass spectrometry detection with electrospray ionisation was evaluated with a QTRAP 6500+ system (SCIEX; Framingham, MA, USA). The lower limit of quantification (LOQ) was 1.5 ng/mL for 8-OHGua and 0.9 ng/mL for 8-OHdG (Cayman Chemical; Ann Arbour, MI, USA). Data acquisition was performed using Analyst®1.7.1 and analysed using SCIEX OS Analytics 1.7.0 software (SCIEX; Framingham, MA, USA). The inter-assay coefficient of variation was low (<10%) for the quality control measures. Levels of OS biomarkers below the LOQ were imputed as the LOQ divided by the square root of 2 (26). Prior to analysis, the urinary OS biomarkers measures were pre-processed to correct for i) time interval between urine collection, processing and storage by fitting a linear model and retaining the residuals (27); ii) batch effects (28); and iii) urine dilution using specific gravity (29). Given the left-skewed distribution, a base-2 log transformation was applied to all OS biomarkers for subsequent analyses (30).

A genetic pathway score for OS (gPFS^ox^) has been previously developed for this cohort (31). In brief, DNA samples were extracted from cord blood and infant 12-month whole blood, and whole-genome genotyping was performed. Twelve genes (four pro-oxidant, eight antioxidant) in a minimal OS pathway were identified. For each of these genes, single-nucleotide genetic polymorphisms (SNPs) linked to gene activity were identified using the GTEx database (32, 33) and the SNP most strongly associated with expression was chosen (non-tissue specific). After assessing the SNPs’ effect on each identified gene (increased or decreased expression), and the impact of each gene on OS (pro- or antioxidant), a cumulative score was calculated for each participant in the BIS. This score reflects the number of pro-oxidant alleles each individual carries for the OS response pathway, and therefore their propensity towards OS (32). The score was divided by 2 to give a score range of 0-12, and participants with a gPFS^ox^ of ≥ 8 (which represented the top 20%) were classified as having a high genetic risk of OS.

### 2.6 Covariates

Several covariates were considered for inclusion in the adjusted model. A directed acyclic graph (DAG) was created based on previous literature and expert knowledge (Supplementary materials, Figure S1). Several of the variables identified had many records with missing data, and multiple imputation was not appropriate given the volume of missing data. Therefore, a minimal set of confounders were included in the models. Postnatal age at lung function was tested as a covariate but was not found to be important in this cohort. Variable selection was predominately decided by the process of the DAG and expert knowledge by the respiratory scientists and physicians on our team, however a formal stepwise regression was performed to test the variable set (including those variables with missing data) which found AIC and R^2^ were better in our selected covariate set, and plots of the residuals showed a better fit when variables with a high volume of missingness were removed. The final model included maternal pre-pregnancy body mass index (BMI), maternal age, maternal prenatal smoking (any vs none), child’s sex and birthweight.

### 2.7 Statistical analysis

Variables were described using histograms and basic statistical summaries. Relationships between variables were explored through plotting and correlation analysis using the Pearson product-moment correlation coefficient. Linear regression was used to explore the association of maternal air pollution exposure with infant lung function at 4 weeks of age, OS biomarkers during pregnancy and inflammation biomarkers during pregnancy (mothers) and from cord blood (child). Modification of the relationship between air pollutants and lung function by presence of an OS genotype in the child was explored through use of interaction terms and stratified analysis. PM_2.5_ and NO_2_ were considered as continuous variables for the primary analyses and results are presented for an IQR increase in pollutant. All analysis was performed in R version 4.2.1, 2022 (R Core Team).

#### 2.7.1 Sensitivity analyses

We performed several sensitivity analyses. Socioeconomic status (SES) was considered as a potential confounder. It has previously been shown in this cohort that inflammation partially mediates the relationship between socioeconomic disadvantage and emotional and behavioural problems in children (25). Education is the socioeconomic indicator recommended by the Organisation for Economic Co-operation and Development for reporting and monitoring socioeconomic inequalities as it is reported with reasonable reliability, can be harmonized across cohorts and countries, generally has few missing data, is relatively stable across adulthood, and is less subject to reverse causality than measures such as income (34, 35). In BIS, maternal education was self-reported in pregnancy in the following categories: < year 10, year 10, year 12, trade, certificate or diploma, bachelor degree or postgraduate degree. Multiplicative interaction terms were added for air pollution and maternal smoking in FRC models. PM_2.5_ and NO_2_ were considered as tertiles, instead of as continuous exposures, to test for non-linearity. Models testing tertiles compared medium to low exposure, and high to low exposure. The effect of air pollution exposure at birth and 1 month of age on lung function was also tested. Lastly, we investigated models that included a child’s exposure to environmental tobacco smoke at 1 month of age as direct exposure to smoke in the home can affect a child’s lung function (36).

## 3 Results

### 3.1 Descriptive results

There were 314 children who had lung function testing conducted at 4 weeks of age and met all the criteria. Of these 51% were male. Maternal smoking during pregnancy was present for 15.6% of children. Mean maternal exposure to PM_2.5_ was 7.5 μg/m^3,^ (min=4.9, max=9.9). Mean maternal NO_2_ was 5.58 ppb (min=1.9, max=15.4). Mean FRC in mL/kg was 18.5, mean LCI was 6.79 lung turnover volumes and tPTEF/tE was 0.37. The cohort summary is presented in Table 1 and Supplementary Table 1.

**Table 1:**
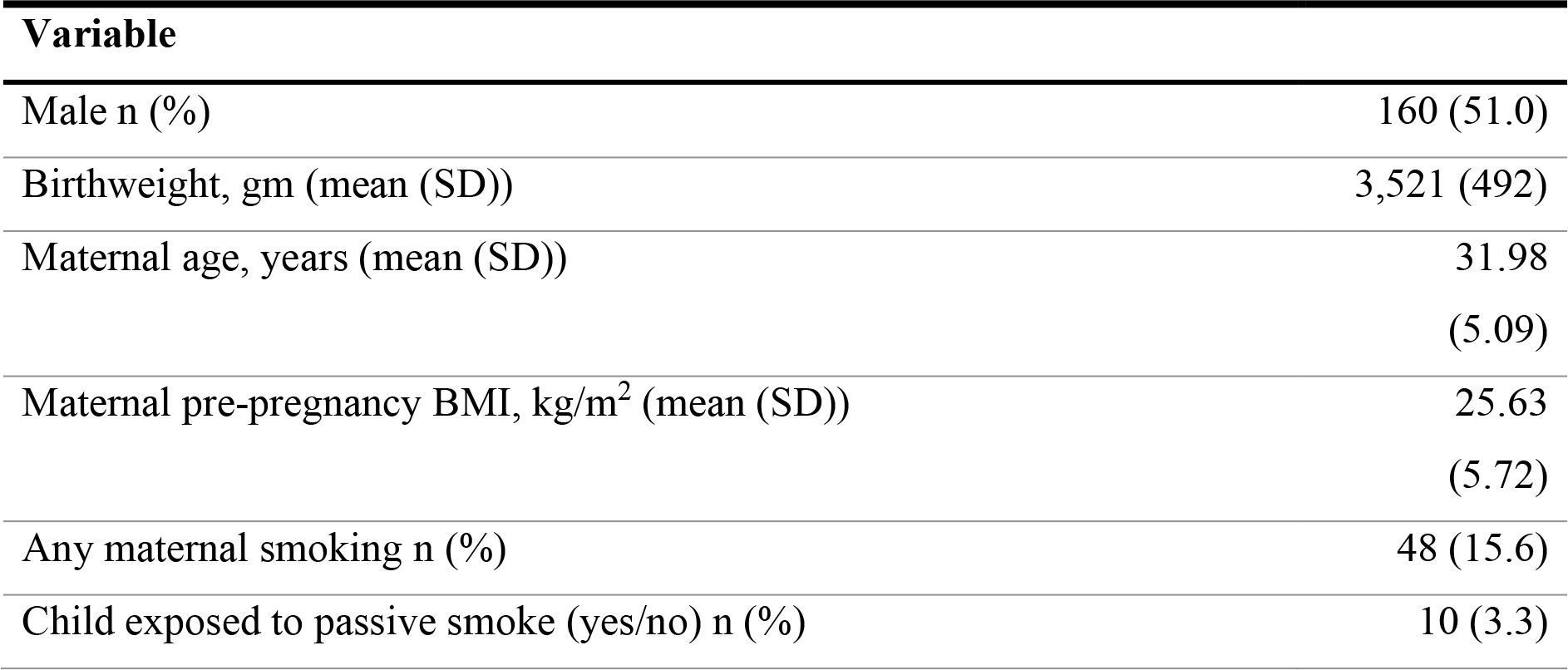
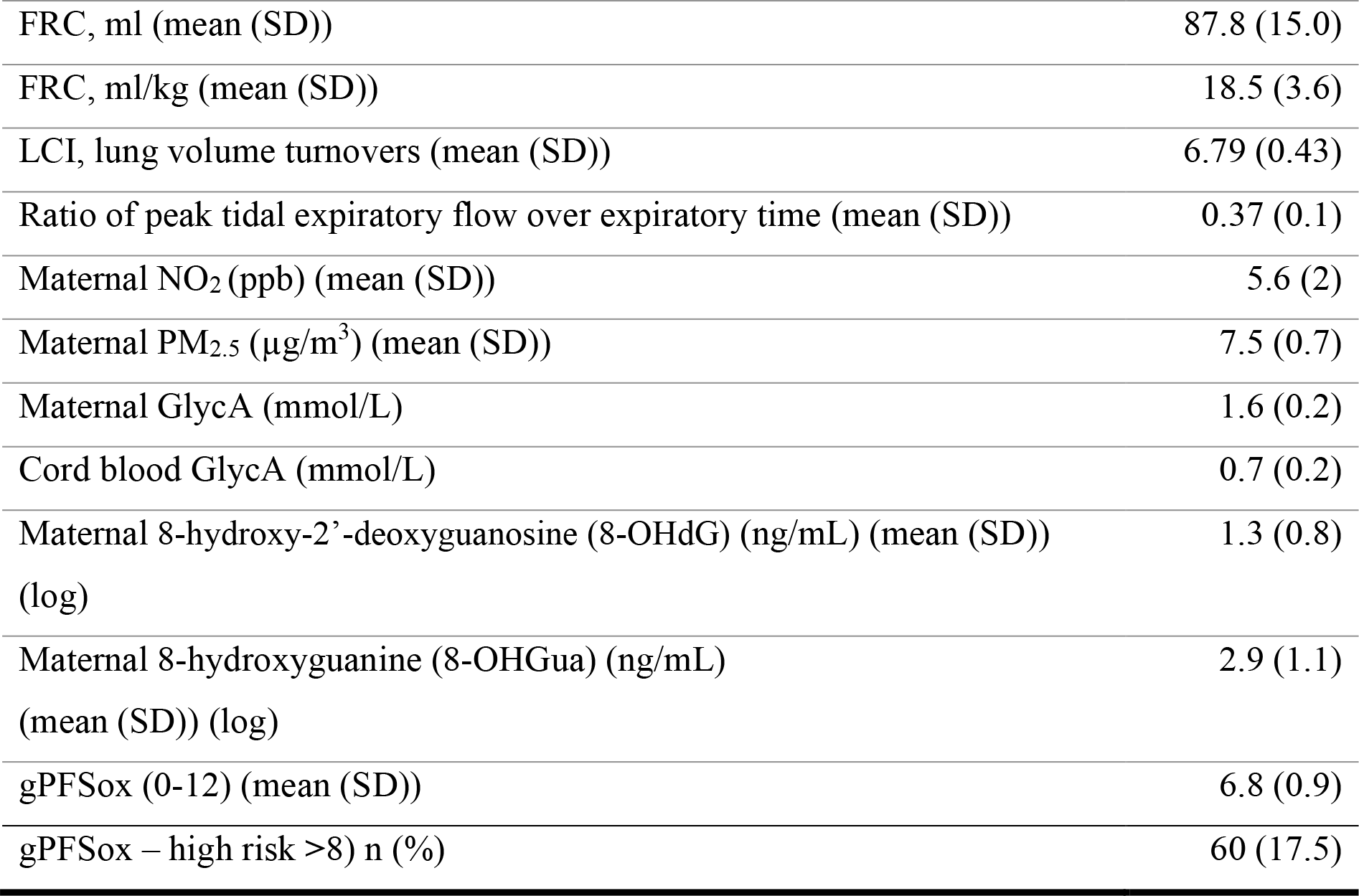
Summary of Barwon Infant Study children and mothers, who completed lung function testing at 4 weeks of age (n=314).

| **Variable** |  |
| --- | --- |
| Male n (%) | 160 (51.0) |
| Birthweight, gm (mean (SD)) | 3,521 (492) |
| Maternal age, years (mean (SD)) | 31.98<br>(5.09) |
| Maternal pre-pregnancy BMI, kg/m<sup>2</sup> (mean (SD)) | 25.63<br>(5.72) |
| Any maternal smoking n (%) | 48 (15.6) |
| Child exposed to passive smoke (yes/no) n (%) | 10 (3.3) |
| FRC, ml (mean (SD)) | 87.8 (15.0) |
| FRC, ml/kg (mean (SD)) | 18.5 (3.6) |
| LCI, lung volume turnovers (mean (SD)) | 6.79 (0.43) |
| Ratio of peak tidal expiratory flow over expiratory time (mean (SD)) | 0.37 (0.1) |
| Maternal NO<sub>2</sub> (ppb) (mean (SD)) | 5.6 (2) |
| Maternal PM<sub>2.5</sub> (µg/m<sup>3</sup>) (mean (SD)) | 7.5 (0.7) |
| Maternal GlycA (mmol/L) | 1.6 (0.2) |
| Cord blood GlycA (mmol/L) | 0.7 (0.2) |
| Maternal 8-hydroxy-2'-deoxyguanosine (8-OHdG) (ng/mL) (mean (SD))<br>(log) | 1.3 (0.8) |
| Maternal 8-hydroxyguanine (8-OHGua) (ng/mL)<br>(mean (SD)) (log) | 2.9 (1.1) |
| gPFSox (0-12) (mean (SD)) | 6.8 (0.9) |
| gPFSox – high risk >8) n (%) | 60 (17.5) |

### 3.2 Air pollution and infant lung function, and modification by genetic propensity to oxidative stress

Table 2 demonstrates the association between air pollution and infant lung function, in the whole population, as well as in children with a genetic propensity to oxidative stress. There was no evidence of a relationship between maternal exposure to air pollution during pregnancy and infant lung function at 4 weeks of age. Maternal exposure to PM_2.5_ was not associated with LCI (β=-0.03 turnovers, 95% CI (−0.09, 0.03), p = 0.34), FRC (β=1.3 mls, 95% CI (−0.9, 3.5), p=0.23) or tPTEF/tE (β=0.003, 95% CI (−0.010, 0.017), p=0.65) at 4 weeks of age in adjusted models. Similarly, maternal exposure to NO_2_ was not associated with LCI (β=0.01 turnovers, 95% CI (−0.04, 0.06), p=0.68), FRC (β=-0.7 mls, 95% CI (−2.5, 1.1), p=0.46) or tPTEF/tE (β=0.003, 95% CI (−0.009, 0.015), p=0.63) at 4 weeks of age in adjusted models.

**Table 2:** Results of linear regression models of maternal exposure to air pollution (IQR increase) and infant lung function at 4 weeks of age in the Barwon Infant Study.

| **Whole cohort** |  |  |  |  |
| --- | --- | --- | --- | --- |
|  |  | *Estimates* | *CI* | *p* |
| **LCI** $n=275$ | PM<sub>2.5</sub> | -0.03 | -0.09, 0.03 | 0.34 |
|  | NO<sub>2</sub> | 0.01 | -0.04, 0.06 | 0.68 |
| **FRC** $n=275$ | PM<sub>2.5</sub> | 1.3 | -0.9, 3.5 | 0.23 |
|  | NO<sub>2</sub> | -0.7 | -2.5, 1.1 | 0.46 |
| **tPTEF/tE** $n=355$ | PM<sub>2.5</sub> | 0.003 | -0.010, 0.017 | 0.65 |
|  | NO<sub>2</sub> | 0.003 | -0.009, 0.015 | 0.63 |
| **Children at high risk of oxidative stress** |  |  |  |  |
|  |  | *Estimates* | *CI* | *p* |
| **LCI** $n=48$ | PM<sub>2.5</sub> | 0.03 | -0.11, 0.17 | 0.67 |
|  | NO<sub>2</sub> | 0.08 | -0.04, 0.20 | 0.18 |
| **FRC** $n=48$ | PM<sub>2.5</sub> | -3.4 | -8.4, 1.5 | 0.18 |
|  | NO<sub>2</sub>* | -5.3 | -9.3, -1.3 | **0.01** |
| **tPTEF/tE** $n=65$ | PM<sub>2.5</sub> | -0.01 | -0.04, 0.02 | 0.49 |
|  | NO<sub>2</sub> | 0.01 | -0.03, 0.03 | 0.99 |
| **Children at low risk of oxidative stress** |  |  |  |  |
|  |  | *Estimates* | *CI* | *p* |
| **LCI** $n=222$ | PM<sub>2.5</sub> | -0.05 | -0.12, 0.03 | 0.23 |
|  | NO<sub>2</sub> | -0.01 | -0.07, 0.06 | 0.84 |
| **FRC** $n=222$ | PM<sub>2.5</sub> | 1.23 | -2.11, 4.57 | 0.47 |
|  | NO<sub>2</sub>* | 0.87 | -1.28, 3.02 | 0.43 |
| **tPTEF/tE** $n=283$ | PM<sub>2.5</sub> | 0.01 | -0.01, 0.02 | 0.24 |
|  | NO<sub>2</sub> | 0.01 | -0.01, 0.02 | 0.41 |
| Models adjusted for sex, birthweight, maternal age, maternal pre-pregnancy BMI and maternal smoking; IQR PM<sub>2.5</sub> = 0.88, IQR NO<sub>2</sub> = 2.27 |  |  |  |  |
| * Difference between estimates for high vs low risk of oxidative stress is $p=0.007$ | | | | |

The relationship between maternal air pollution exposure and lung function was explored in the context of a child’s genetic risk for OS. We found evidence of an association between NO_2_ and reductions in lung function for those children with a gPFSox indicating a high risk of OS. For an IQR increase (ppb) of NO_2_ there was a 5.3 mL decrease in FRC at 4 weeks of age (β=-5.3 mls, 95% CI (−9.3, -1.3), p=0.01). An IQR increase in maternal NO_2_ was not associated with LCI (β=0.08 turnovers, 95% CI (−0.04, 0.20), p=0.18). However, there is a potential for a threshold response.

We did not find a relationship between maternal exposure to PM_2.5_ and either FRC (β=-3.4 mls, 95% CI (−8.4, 1.5), p=0.18) or LCI (β=0.03 turnovers, 95% CI (−0.11, 0.17), p=0.67) LCI in children at high risk of OS. This did not change when using exposure in tertiles (see table S8 in table). Finally, there was no association with either PM_2.5_ (β -0.01, 95% CI (−0.04, 0.02), p=0.49) or NO_2_ (β 0.01, 95% CI (−0.03, 0.03), p=0.99) and tPTEF/tE at 4 weeks of age, in children at higher risk of OS.

### 3.3 Air pollution and inflammation

Table 3 shows the estimated effect of air pollution on GlycA, a marker of chronic inflammation, in mothers and in cord blood. There was no relationship between maternal exposure to PM_2.5_ (β=-0.01 mmol/l, 95% CI (−0.02, 0.01), p=0.50) or NO_2_ (β=0.003 mmol/l, 95% CI (−0.012, 0.017), p=0.73) and GlycA in maternal blood in fully adjusted models. Further, there was no relationship between maternal exposure to PM_2.5_ (β=0.01 mmol/l, 95% CI (−0.02, 0.04), p=0.56) or NO_2_ (β=0.05 mmol/l, 95% CI (−0.017, 0.028), p=0.64) and GlycA in cord blood in fully adjusted models.

**Table 3:** Results of linear regression models of maternal exposure to air pollution and maternal and child oxidative stress biomarkers and inflammation in the Barwon Infant Study.

|  | Air pollutants | Estimates | CI | p |
| --- | --- | --- | --- | --- |
| Oxidative stress biomarkers<sup>#</sup> |  |  |  |  |
| Maternal 8-hydroxy-2'-deoxyguanosine<sup>^</sup> | PM<sub>2.5</sub> | 0.08 | -0.07, 0.21 | 0.33 |
|  | NO<sub>2</sub> | 0.01 | -0.12, 0.13 | 0.93 |
| Maternal 8-hydroxyguanine<sup>^</sup> | PM<sub>2.5</sub> | 0.02 | -0.16, 0.21 | 0.81 |
|  | NO<sub>2</sub> | 0.03 | -0.13, 0.19 | 0.69 |
| Inflammation biomarker |  |  |  |  |
| Maternal GlycA<sup>^</sup> | PM<sub>2.5</sub> | -0.01 | -0.02, 0.01 | 0.50 |
|  | NO<sub>2</sub> | 0.003 | -0.012, 0.017 | 0.73 |
| Cord blood GlycA<sup>*</sup> | PM<sub>2.5</sub> | 0.01 | -0.02, 0.04 | 0.56 |
|  | NO<sub>2</sub> | 0.05 | -0.017, 0.028 | 0.64 |
| <sup>#</sup>Oxidative stress biomarkers were pre-processed for time interval between urine collection, processing and storage, batch effects, and urine dilution using specific gravity.<br><sup>^</sup>Models adjusted for maternal age, maternal pre-pregnancy BMI and maternal smoking<br><sup>*</sup>Models adjusted for sex, birthweight, maternal age, maternal pre-pregnancy BMI and maternal smoking |  |  |  |  |

### 3.4 Air pollution and oxidative stress

Table 3 demonstrates the association between air pollution and maternal oxidative stress. There was no relationship between maternal exposure to air pollution and biomarkers for OS during pregnancy or in cord blood. There was no association between PM_2.5_ and urinary 8-OHdG (β=0.08 ng/mL, 95% CI -0.07, 0.21, p=0.33) or NO_2_ (β=0.01 ng/mL, 95% CI (−0.12, 0.13), p=0.93) in fully adjusted models. The same was true of urinary 8-OHGua, which was not associated with PM_2.5_ (β=0.02 ng/mL, 95% CI (−0.16, 0.21), p=0.81) nor NO_2_ (β=0.03 ng/mL, 95% CI (−0.13, 0.19), p=0.69) in adjusted models.

### 3.5 Sensitivity analysis

Several sensitivity analyses were performed: using a child’s own passive smoking exposure, instead of maternal smoking, as a covariate; an interaction term between air pollution and maternal smoking; air pollutants as tertiles; the effect of SES; and a child’s direct exposure to air pollution and lung function. There were no significant associations in any of our sensitivity analyses and model fit was not improved compared with the main models, with the exception of high exposure to NO_2_ and lung function measures. When including NO_2_ in tertiles, the highest tertile of NO_2_ was associated with differences in both FRC (β=-13.1 mls, 95% CI (−26.1, -0.1), p=0.05) and LCI (β=0.46 turnovers, (95% CI 0.10, 0.82), p=0.01) in children with a genetic propensity to oxidative stress (see table S7 in supplement, difference in effect from children at low risk of OS is *p=*0.02 and *p=*0.007, respectively). (See supplementary materials). A sensitivity analysis was performed using an interaction term between a child’s gPFSox and maternal air pollution for each lung function outcome, but none were significant (data not shown).

## 4 Discussion

Our study found that high levels of exposure to NO_2_ is associated with lower FRC and higher LCI in infants with a genetic propensity to oxidative stress. There was no relationship between maternal exposure to air pollution and infant lung function, in the whole population. We found no overall association between air pollution and either maternal or cord blood inflammation biomarkers. Our results are largely in keeping with those of Latzin et al. (10), who did not find an association with MBW parameters, although they did report an association between PM_10_ and tidal breathing flows. We did not have data on PM_10_ to assess the effect of the larger size fraction in our cohort. Decrue et al (9) reported an association between minute ventilation and maternal exposure to PM_10_ and NO_2_ in the second trimester, especially in preterm infants, however they did not find a relationship with tPTEF/tE and either air pollutant. In our study, there were only 15 participants born preterm (< 37 weeks gestational age) with lung function data, and univariate analysis did not find an association between air pollution exposure in the antenatal period and lung function at 4 weeks in preterm infants (data not shown).

The lack of association between air pollution and OS was surprising. Our sample size is modest for testing associations with air pollution, which tend to have small effect estimates (37), which is an issue in post hoc analysis where the cohort is powered for clinical outcomes. The Barwon Infant Study was designed to minimise participant burden while achieving deep phenotyping, but it has previously being recognised that this may come at a cost of statistical power, especially for uncommon outcomes and those with small effect sizes (15). It has previously been found that circulating biomarkers are less sensitive to air pollution exposure than more localised lung measures. Zhang et al (38) found an association between various air pollutants and malondialdehyde in expired breath condensate, however they found little relationship between air pollutants and oxidised low density lipoprotein or interleukin-6.

Future research on this topic should focus on respiratory-specific markers of oxidation, such as malondialdehyde (11, 12) or glutathione sulfonamide (39). Previous work in this cohort found that a high risk OS genotype combined with higher exposure to phthalates in utero was associated with an elevated risk of adverse neurodevelopmental outcomes when compared with children who had a genotype not at increased risk of OS (40). It is possible that lung function development is protecting by the buffering and defence mechanisms occurring in utero, especially the protection offered by the placenta. There is a well developed body of literature and meta-analysis showing that endocrine disrupting chemicals cross the placenta, and affect the development of key systems (41, 42). However, it is less clear how air pollutants influence the development of children, and which systems are vulnerable. This is an area that requires more research. Lastly, we need to consider the level of air pollution exposure in this cohort, and whether the pollution levels are too low to impact on lung development. This is unlikely given the previous studies were also conducted in communities with historically good air quality. We hypothesized that children with a genotype predisposing them to a higher risk of OS would have a stronger negative association between maternal air pollution exposure and infant lung function, which was true for NO_2_. This indicates that examining the impact of air pollution on those with genetic vulnerability such as those with a low genetic antioxidant capacity is informative.

The Barwon Infant Study is a unique and data rich cohort, that affords our study several methodological strengths. Lung function testing was performed in non-sedated infants and followed international guidelines (22). Participants were recruited prospectively in the prenatal period from all eligible members of the population, and therefore there in no selection bias in regard to exposure or outcome. Our air pollution model is externally validated and has been used in previous studies (17, 18). Urinary oxidative stress biomarkers were measured using the ‘gold-standard’ approach of LC-MS/MS and pre-processed to reduce the number of variables required in the model. However, limitations remain. The sample size is modest, as discussed earlier. Not all parents consented to lung function testing, and only 55% of infants who attempted lung function returned acceptable and reproducible results. Our air pollution model is limited to annual averages, which does not allow us to measure short-term temporal changes or trimester specific effects. We only have a single time point for the OS biomarkers. We did not have OS biomarkers in the infants. Lastly, our sample was not large enough for sex-specific analysis and may be subject to unmeasured confounding.

## 5 Conclusion

Our study finds no association between maternal exposure to PM_2.5_ or NO_2_ in the prenatal period and infant lung function at four weeks of age, with the exception of NO_2_ for children at higher genetic risk of OS. Further, we did not find an association between maternal exposure to air pollution and OS biomarkers in maternal blood or cord blood.

## Data Availability

All data produced in the present study are only available under strict approval conditions for approved research projects from the Barwon Infant Study

## Acknowledgements

The authors thank the *Barwon Infant Study* participants for their valuable contribution. The establishment work and infrastructure for the BIS was provided by the Murdoch Children’s Research Institute, Deakin University and Barwon Health. Subsequent funding was secured from the National Health and Medical Research Council of Australia, The Jack Brockhoff Foundation, the Scobie Trust, the Shane O’Brien Memorial Asthma Foundation, the Our Women’s Our Children’s FundRaising Committee Barwon Health, The Shepherd Foundation, the Rotary Club of Geelong, the Ilhan Food Allergy Foundation, GMHBA Limited and the Percy Baxter Charitable Trust, Perpetual Trustees, and the Minderoo Foundation. In-kind support was provided by the Cotton On Foundation and CreativeForce. Research at Murdoch Children’s Research Institute is supported by the Victorian Government’s Operational Infrastructure Support Program. C Pham is supported by a Melbourne Children’s Campus LifeCourse PhD Support Program scholarship, funded by Royal Children’s Hospital Foundation grant number 2018-984. This work was also supported by NHMRC, Australia Investigator Grants [APP1197234 to AL Ponsonby]. We thank Terry Dwyer for his role in the original BIS Steering Committee. The BIS Investigator Group consists of John Carlin, Mimi Tang, Len Harrison, Fiona Collier, Amy Loughman, Sarath Ranganathan and Lawrence Gray. D Vilcins was supported by the Louisiana State University Superfund Research program and the National Institute Of Environmental Health Sciences of the National Institutes of Health under Award Number P42ES013648. The content is solely the responsibility of the authors and does not necessarily represent the official views of the National Institutes of Health.

D Burgner is supported by a National Health and Medical Research Council (Australia) Investigator Grant APP1175744. Research at Murdoch Children’s Research Institute is supported by the Victorian Government’s Operational Infrastructure Support Program.

