## Supplement for "Association of maternal air pollution exposure and infant lung function is modified by genetic propensity to oxidative stress"

**Affiliations:**

### Supplementary materials

#### 1 Directed acyclic graph

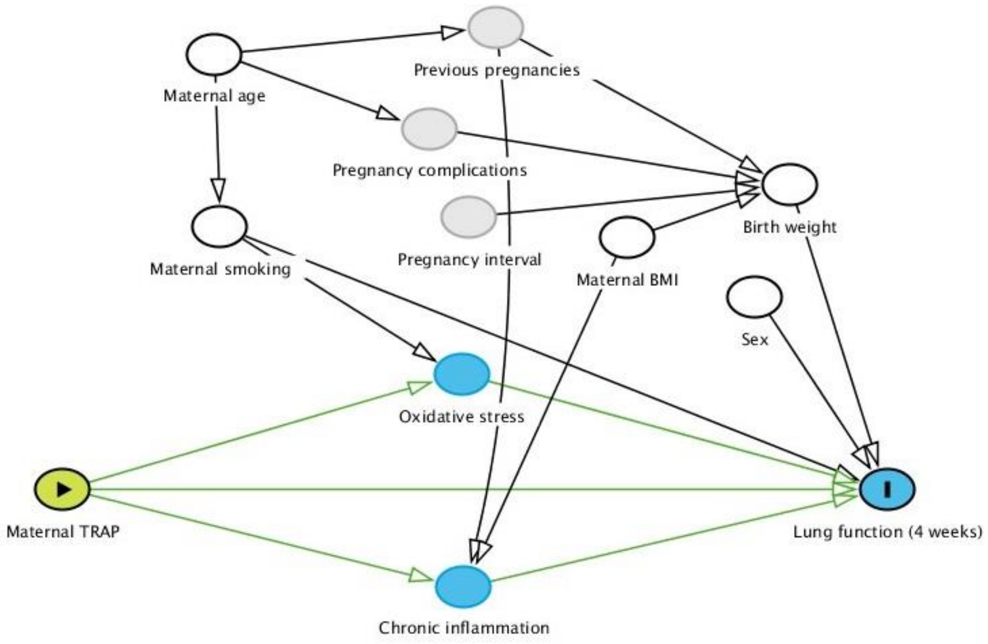


Figure 1: Directed acyclic graph depicted proposed relationships exposure, outcome and covariates. Green shapes and arrows indicate exposure, blue shapes indicate outcome and its antecedents, including potential mediators. White shapes indicate variables which were included as covariates, while grey shapes are variables proposed to have a relationship on the causal network, but which are unobserved in our models. Maternal TRAP refers to maternal exposure to traffic related air pollution.

### 2 Summary of exposure, outcome and mediators

Table S1: Summary of exposures and outcomes in the Barwon Infant Study

| Variable | mean | sd | median | min | max | IQR |
|---|---|---|---|---|---|---|
| FRC (mL / kg) | 87.7 | 14.9 | 86.7 | 52.6 | 144.0 | 4.27 |
| LCI (turnovers) | 6.79 | 0.43 | 6.79 | 5.410 | 8.2 | 0.52 |
| Maternal $NO_2$ (ppb) | 5.58 | 1.97 | 5.44 | 1.983 | 13.9 | 2.27 |
| Maternal $PM_{2.5}$ (μg/$m^3$) | 7.50 | 0.66 | 7.42 | 5.932 | 9.9 | 0.88 |
| Maternal 8-OHdG (ng/mL) (log) | 1.3 | 0.8 | 1.3 | -1.3 | 4.1 | 0.94 |
| Maternal 8-OHGua (ng/mL) (log) | 2.9 | 1.1 | 3.0 | -0.7 | 6.5 | 1.09 |
| gPFSox | 6.8 | 0.9 | 7.00 | 4.5 | 9.5 | 1.5 |

### 3 Sensitivity analysis

#### 3.1 SES

Adding socio-economic status to the model (using maternal education level as a proxy measure) of FRC did not significantly change any of the estimates and was not itself significant (β=0.6 mls, 95% CI (-1.7, 2.8), p=0.63).

#### 3.2 Air pollution and maternal smoking interaction (adjusted models)

Table S2: Estimates for lung function at 4 weeks of age (FRC) for maternal exposure to $PM_{2.5}$ and the interaction with maternal smoking in the Barwon Infant Study

| | **median FRC** | | |
|---|---|---|---|
| *Predictors* | *Estimates* | *CI* | *p* |
| Maternal $PM_{2.5}$ in year before birth (ug/m3) | 1.2 | -1.5 – 4.0 | 0.374 |
| $PM_{2.5}$ * maternal smoking | 1.6 | -5.2 – 8.3 | 0.650 |
| Models adjusted for sex, birthweight, maternal age, maternal pre-pregnancy BMI and maternal smoking | | | |

#### 3.3 Child's passive smoke exposure (adjusted)

Table S3: Estimates for lung function at 4 weeks of age (LCI) for maternal exposure to $PM_{2.5}$, maternal smoking, and a child's exposure to environmental tobacco smoke at 4 weeks of age in the Barwon Infant Study

| | **median LCI** | | |
|---|---|---|---|
| *Predictors* | *Estimates* | *CI* | *p* |
| $PM_{2.5}$ (ug/m3) | -0.03 | -0.10 – 0.05 | 0.446 |
| Maternal smoking | -0.04 | -0.17 – 0.10 | 0.612 |
| Child passive smoke exposure | 0.01 | -0.29 – 0.31 | 0.947 |

Models adjusted for sex, birthweight, maternal age, maternal pre-pregnancy BMI and maternal smoking

Table S4: Estimates for lung function at 4 weeks of age (LCI) for maternal exposure to $NO_2$, maternal smoking, and a child's exposure to environmental tobacco smoke at 4 weeks of age in the Barwon Infant Study

| | **median LCI** | | |
|---|---|---|---|
| *Predictors* | *Estimates* | *CI* | *p* |
| $NO_2$ (ppb) | 0.01 | -0.02 – 0.03 | 0.580 |
| Maternal smoking | -0.03 | -0.17 – 0.10 | 0.619 |
| Child passive smoke exposure | 0.02 | -0.27 – 0.32 | 0.871 |

Models adjusted for sex, birthweight, maternal age, maternal pre-pregnancy BMI and maternal smoking

#### 3.4 Child's exposure to $PM_{2.5}$ and lung function (adjusted)

Table S5: Effect estimates for lung function at 4 weeks of age (FRC) for a child's exposure to $PM_{2.5}$ at different timepoints, including an interaction with the oxidative stress genotype, in the Barwon Infant Study.

| | median FRC | | |
|---|---|---|---|
| *Predictors* | *Estimates* | *CI* | *p* |
| $PM_{2.5}$ (mcg/m3) at birth | 1.2 | -1.3 – 3.6 | 0.339 |
| $PM_{2.5}$ mcg/m3) at 1 month | 0.6 | -1.8 – 3.0 | 0.596 |
| $PM_{2.5}$ mcg/m3) at 1 month * oxidative stress genotype | 4.1 | -1.9 – 10.0 | 0.178 |
| Models adjusted for sex, birthweight, maternal age, maternal pre-pregnancy BMI and maternal smoking | | | |

Table S6: Effect estimates for lung function at 4 weeks of age (FRC) for a child's exposure to $NO_2$ at different timepoints, including an interaction with the oxidative stress genotype, in the Barwon Infant Study.

| | median LCI | | |
|---|---|---|---|
| *Predictors* | *Estimates* | *CI* | *p* |
| $PM_{2.5}$ (mcg/m3) at birth | 0.002 | -0.069 – 0.072 | 0.959 |
| $PM_{2.5}$ (mcg/m3) at 1 month | 0.012 | -0.058 – 0.081 | 0.742 |
| $PM_{2.5}$ (mcg/m3) at 1 month * oxidative stress genotype | -0.073 | -0.246 – 0.099 | 0.404 |
| Models adjusted for sex, birthweight, maternal age, maternal pre-pregnancy BMI and maternal smoking | | | |

#### 3.5 Air pollutants in tertiles

Table S7: Results of linear regression models of maternal exposure to air pollution and infant lung function at 4 weeks of age in the Barwon Infant Study, sensitivity using air pollution variable in tertiles

| **Whole cohort** | | | | |
|---|---|---|---|---|
| | | *Estimates* | *CI* | *p* |
| **LCI** *n*=275 | Medium $PM_{2.5}$ | -0.02 | -0.14 – 0.10 | 0.78 |
| | High $PM_{2.5}$ | -0.02 | -0.14 – 0.10 | 0.73 |
| | Medium $NO_2$ | -0.09 | -0.21 – 0.03 | 0.13 |
| | High $NO_2$ | -0.01 | -0.13 – 0.11 | 0.91 |
| **FRC** *n*=275 | Medium $PM_{2.5}$ | 1.33 | -2.81 – 5.47 | 0.53 |
| | High $PM_{2.5}$ | 3.14 | -1.04 – 7.32 | 0.14 |
| | Medium $NO_2$ | 2.58 | -1.58 – 6.73 | 0.22 |
| | High $NO_2$ | 1.34 | -2.85 – 5.53 | 0.53 |
| **tPTEF/tE** *n*=355 | Medium $PM_{2.5}$ | -0.01 | -0.03 – 0.02 | 0.47 |
| | High $PM_{2.5}$ | 0.00 | -0.02 – 0.03 | 0.92 |
| | Medium $NO_2$ | -0.02 | -0.05 – 0.00 | 0.10 |
| | High $NO_2$ | 0.001 | -0.03 – 0.03 | 0.97 |
| **Children at high risk of oxidative stress** | | | | |
| | | *Estimates* | *CI* | *p* |
| **LCI** *n*=48 | Medium $PM_{2.5}$ | 0.14 | -0.17 – 0.46 | 0.36 |
| | High $PM_{2.5}$ | -0.02 | -0.34 – 0.30 | 0.91 |
| | Medium $NO_2$ | 0.22 | -0.11 – 0.54 | 0.19 |
| | High $NO_2$* | 0.46 | 0.10 – 0.82 | **0.01** |
| **FRC** *n*=48 | Medium $PM_{2.5}$ | 0.6 | -11.0 – 12.1 | 0.92 |
| | High $PM_{2.5}$ | -0.6 | -12.2 – 11.1 | 0.92 |
| | Medium $NO_2$ | -0.9 | -12.6 – 10.8 | 0.88 |
| | High $NO_2$^ | -13.1 | -26.1 – -0.1 | **0.05** |
| **tPTEF/tE** *n*=65 | Medium $PM_{2.5}$ | -0.04 | -0.10 – 0.03 | 0.31 |
| | High $PM_{2.5}$ | -0.02 | -0.09 – 0.05 | 0.64 |
| | Medium $NO_2$ | -0.03 | -0.10 – 0.03 | 0.33 |
| | High $NO_2$ | 0.02 | -0.05 – 0.09 | 0.51 |

Models adjusted for sex, birthweight, maternal age, maternal pre-pregnancy BMI and maternal smoking
* Estimates significantly different to model of low risk OS kids *p*=0.007
^ Estimates significantly different to model of low risk OS kids *p*=0.02
